# Genetically predicted 25-Hydroxyvitamin D levels on Hypothyroidism: A two-sample Mendelian Randomization

**DOI:** 10.1101/2023.08.30.23294811

**Authors:** Mahdi Akbarzadeh, Sahand Tehrani Fateh, Aysan Moeinafshar, Danial Habibi, Amir Hossein Ghanooni, Amir Hesam Saeidian, Parisa Riahi, Maryam Zarkesh, Hossein Lanjanian, Mina Jahangiri, Maryam Moazzam-Jazi, Farshad Teymoori, Fereidoun Azizi, Mehdi Hedayati, Maryam Sadat Daneshpour

## Abstract

**Background:** Alterations in levels of 25-Hydroxyvitamin D have been associated with the risk of thyroid disease. This study uses Mendelian randomization (MR) to infer the possible causal association of 25-Hydroxyvitamin D with hypothyroidism.

**Methods:** We performed two-sample MR using the summary statistics data from genome-wide association studies (GWAS) from populations with European ancestry to infer the causality of genetically controlled levels of 25-Hydroxyvitamin D on the risk of hypothyroidism, Hashimoto’s thyroiditis, as well as biochemical parameters of thyroid diseases. The inverse-variance method (IVW) was used as the primary method to calculate the combined effect of all SNPs. Other methods were adopted to evaluate the stability and reliability of the results. Comprehensive sensitivity analyses were conducted to ensure that none of the MR analysis’s primary assumptions were violated.

**Results:** The results of the IVW analysis revealed a significant causal association between higher levels of 25-Hydroxyvitamin D and lower risk of hypothyroidism (beta = −0.197, 95% CI (− 0.301, −0.093); SE = 0.053, P_beta_ = 2.256×10^-4^) as well as increased levels of free T4 (beta = 0.204, 95% CI (0.305, 0.094); SE = 0.056, P_beta_ = 3.0506×10^−4^). On the other hand, no significant causality was determined for higher levels of 25-Hydroxyvitamin D in association with Hashimoto’s thyroiditis (beta=-0.047, 95% CI (−0.245, 0.151), p=0.641) and TSH levels (beta = −0.024, 95% CI (−0.099, - 0.051); P_beta_ = 0.524).

**Conclusion:** The results of this two-sample MR study provide evidence supporting the potential of 25-Hydroxyvitamin D supplementation in reducing the risk of hypothyroidism.

## Introduction

Hypothyroidism is a condition characterized by increased serum levels of thyroid-stimulating hormone (TSH), which can be accompanied by a reduction of free T4 (fT4) levels in cases of overt hypothyroidism, in contrast to its normal levels in the subclinical condition (1). The prevalence of the overt form of this condition has been estimated between 0-2% and 3-5% in the European populations, and higher prevalence has been reported among female sex, elderly (>65 years), and Caucasian populations. In addition, hypothyroidism has been observed as a comorbidity of several autoimmune diseases, including type 1 diabetes and celiac disease. On the contrary, cigarette smoking and moderate levels of alcohol consumption have been considered protective factors against hypothyroidism (2). Both iodine deficiency and excess, especially by consumption of iodine-containing drugs and radioiodine treatment, can cause primary hypothyroidism, and iodine deficiency is the leading cause of this condition (3–7). The most common cause of hypothyroidism in iodine-sufficient regions is Hashimoto thyroiditis (2). Central and peripheral hypothyroidism, associated with dysfunction of the pituitary/hypothalamus and deiodinase three enzymes, respectively, are among the relatively rare causes of hypothyroidism (8, 9).

To date, a variety of genetic and environmental factors have shown a significant association with the incidence of autoimmune thyroid diseases. History of smoking, alcohol consumption, stress, infectious diseases, pharmacological agents, increased levels of iodine and selenium, and 25- Hydroxyvitamin D deficiency are among such environmental factors (10). 25-Hydroxyvitamin D is an important mediator of calcium and phosphorus homeostasis, which its role has also been implicated in a variety of systemic diseases, including diabetes, cardiovascular diseases, metabolic syndrome, and autoimmunity (11–13).

Many studies have shown a significant association between lower 25-Hydroxyvitamin D levels, hypothyroidism, and increased TSH levels. The association may, in turn, be due to the similarity between the 25-Hydroxyvitamin D receptor (VDR) and the nuclear thyroid hormone receptors and 25-Hydroxyvitamin D’s ability to regulate the secretion of TSH (14). 25-Hydroxyvitamin D has also been identified as an immunomodulatory agent, and VDR is expressed in the majority of immune cells, which is involved in the regulation of proliferation, differentiation, and phagocytosis-related processes of these cell subsets in response to its ligand, 25-Hydroxyvitamin D (15). Several studies have shown that dysregulation in such inflammatory processes due to 25- Hydroxyvitamin D deficiency can be associated with the pathogenesis of Graves’ disease and Hashimoto’s thyroiditis (16, 17).

The causal association between 25-Hydroxyvitamin D levels and hypothyroidism has not been well investigated in the literature. A limited number of randomized controlled trials have been conducted on the effect of 25-Hydroxyvitamin D supplementation on biochemical characteristics of cases of hypothyroidism(14, 18). Due to the limitations of this type of study, an alternative approach can be used to assess such causal relationships.

Mendelian randomization (MR), as a beneficial and growingly favored epidemiological method, exploits genome-wide association (GWAS) summary statistics to infer causal relationships between specific exposure and outcome variables by using genetic variants as instrumental variables (IVs) without facing the limitations of randomized control trials and confounding variables of the observational studies (19).

In this study, as the first study of its kind, we have conducted an MR analysis based on the latest GWAS findings worldwide, including more than 400k participants, to determine the causal relationship between 25-Hydroxyvitamin D and hypothyroidism among the European population.

## Method

### Study Design

MR analysis was employed to investigate the causal relationship between 25-Hydroxyvitamin D levels in serum and hypothyroidism. Hypothyroidism as a general condition, Hashimoto’s thyroiditis as a type of autoimmune hypothyroidism, T4 and TSH as the markers of hypothyroidism were assessed in this study.

The summary-level data for conducting MR analysis were obtained from the GWAS catalog database, namely hypothyroidism (GCST90204167), Hashimoto’s thyroiditis (GCST90018855), and 25-Hydroxyvitamin D (GCST90000616). The summary-level data for TSH and fT4 was also taken from the Thyroid Omics Consortium. The Details of summary-level data for each GWAS are available in Table 1.

**Table 1.**
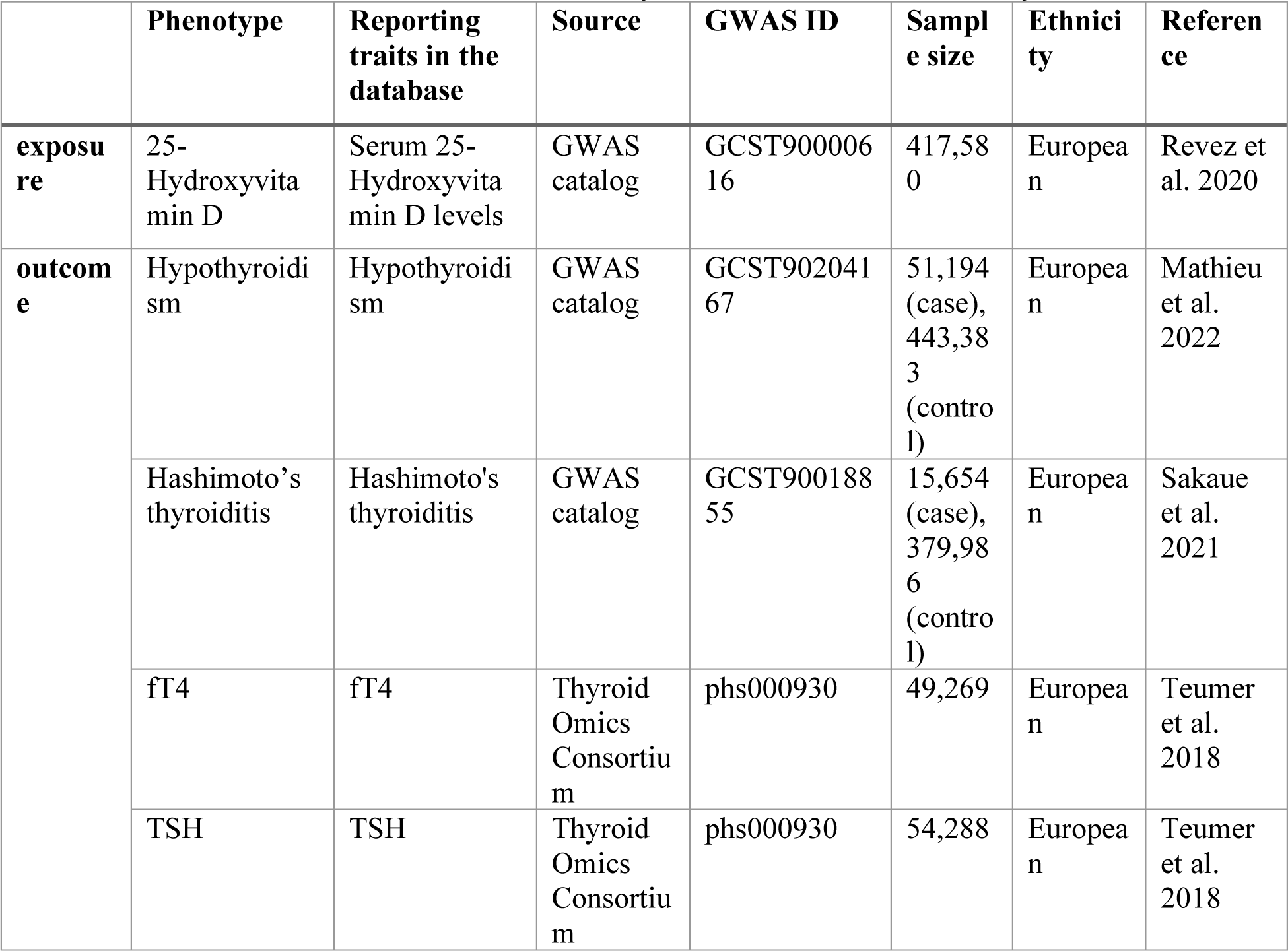
Detailed information of GWAS summary statistics data used for analyses.

|  | Phenotype | Reporting traits in the database | Source | GWAS ID | Sample size | Ethnicity | Reference |
| --- | --- | --- | --- | --- | --- | --- | --- |
| **exposure** | 25-Hydroxyvitamin D | Serum 25-Hydroxyvitamin D levels | GWAS catalog | GCST90000616 | 417,580 | European | Revez et al. 2020 |
| **outcome** | Hypothyroidism | Hypothyroidism | GWAS catalog | GCST90204167 | 51,194 (case), 443,383 (control) | European | Mathieu et al. 2022 |
|  | Hashimoto's thyroiditis | Hashimoto's thyroiditis | GWAS catalog | GCST90018855 | 15,654 (case), 379,986 (control) | European | Sakaue et al. 2021 |
|  | ft4 | ft4 | Thyroid Omics Consortium | phs000930 | 49,269 | European | Teumer et al. 2018 |
|  | TSH | TSH | Thyroid Omics Consortium | phs000930 | 54,288 | European | Teumer et al. 2018 |

### Assumptions of MR study

In this study, three main assumptions are considered for MR analysis: 1) the SNPs selected as instrumental variables (IVs) should be strongly associated with the 25-Hydroxyvitamin D levels; 2) the IVs must not have a relationship with the confounders of 25-Hydroxyvitamin D levels and hypothyroidism association; and 3) the IVs should affect hypothyroidism merely through 25- Hydroxyvitamin D levels and do not have a direct association with hypothyroidism (20) (Figure 1). The validity of these assumptions should be tested in every MR analysis.

**Figure 1.**
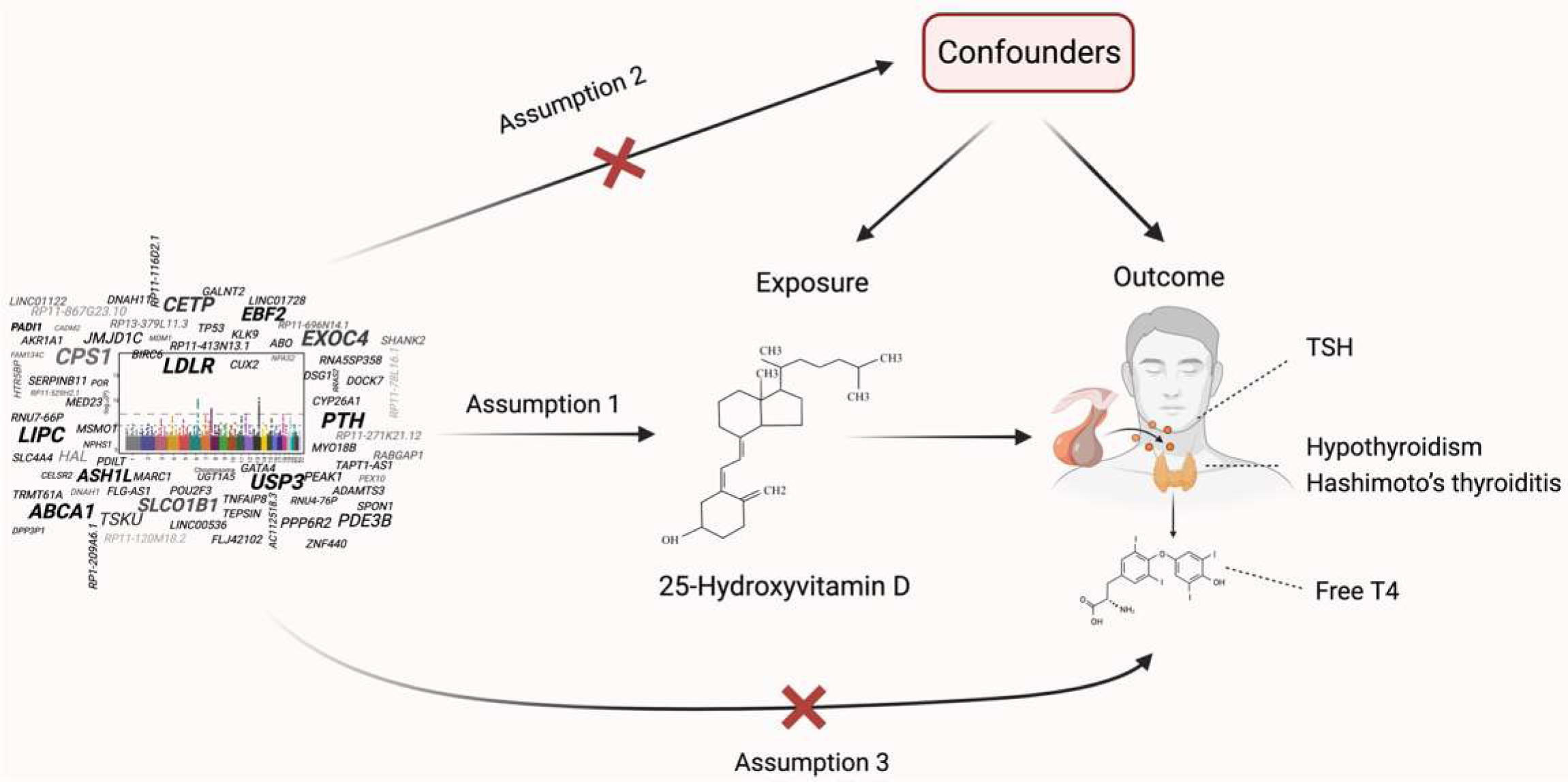
Directed acyclic graph of the MR framework investigating the causal relationship between 25- Hydroxyvitamin D levels and Hypothyroidism. The main assumptions of MR analysis 1) the SNPs selected as instrumental variables (IVs) should be strongly associated with the 25-Hydroxyvitamin D levels; 2) the IVs must not have a relationship with the confounders of 25-Hydroxyvitamin D levels and hypothyroidism association; and 3) the IVs should affect hypothyroidism merely through 25-Hydroxyvitamin D levels and do not have a direct association with hypothyroidism

### Selection of instrumental variables (IVs)

The two-sample MR was conducted using genetic data extracted from GWAS summary statistics. *TwoSampleMR* package in R was employed to integrate and analyze the data. SNPs associated with 25-Hydroxyvitamin D levels in serum that passed the genome-wide significance threshold (P < 5×10-8) were selected as IVs and then were clumped (r^2^ ≤ 0.001; clumping window, 10,000 kb) to ensure the independence of selected SNPs. We harmonized the summary data of 25- Hydroxyvitamin D and hypothyroidism, Hashimoto’s thyroiditis, fT4, and TSH to check whether alleles are genuinely aligned and also to exclude palindromic SNPs if they exist. The strength of each IV was assessed using the F-statistics (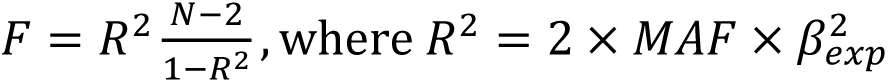, and *R*^2^ is the extracted SNPs’ total variance, and N is the sample size) and IV with F-statistics lower than 10 were excluded.

### Two-sample MR analysis

The inverse variance-weighted (IVW) method was used as the primary method to calculate the combined effect of all SNP s. A P-value below 0.0125 (0.05/4, 4 is the number of outcomes) was considered statistically significant after the Bonferroni correction. Additionally, MR Egger, Penalized MR-Egger, Robust MR-Egger, Penalized robust MR-Egger, Simple Median, Weighted Median, Penalized Weighted Median, Simple Mode, Weighted Mode, Penalized IVW, Robust IVW, Penalized robust IVW, MR Lasso, MR-cML, Debiased inverse-variance weighted, and mode-based estimation (MBE) method were adopted to evaluate the stability and reliability of results (21). We conducted a comprehensive sensitivity analysis to ensure that none of the MR analysis primary assumptions were violated. Two-sample MR is prone to heterogeneity due to differences in analysis platforms, populations, and experimental conditions.

Regarding heterogeneity, we used Cochran’s Q statistic and the I^2^ index for MR-inverse-variance weighted analyses and Rucker’s Q statistic for MR-Egger to detect heterogeneity (22, 23). Funnel plot was also used to visualize the data and detect the outlier SNPs. Moreover, we used the MR-Egger method (by intercept tests) to evaluate horizontal pleiotropy (23). We also searched the human gene phenotypic association database (phenoscanner V2) (http://www.phenoscanner.medschl.cam.ac.uk) and removed pleiotropic SNPs directly associated with outcome or confounder variables. The Mendelian Randomization Pleiotropy RESidual Sum and Outlier (MR-PRESSO) test was performed to detect possible outliers, correcting for horizontal pleiotropy via outlier removal (24).

Cook’s distance was used to diagnose and eliminate SNPs that strongly and falsely influenced the fitted values of the model. SNPs detected as outliers and influential points were excluded for further MR analysis. Leave-one-out and single SNP MR were used as sensitivity analyses (25). All the statistical analyses were performed in R software 4.3.1 implemented in R Studio using “*TwoSampleMR*”, “*MendelianRandomization*”, “*MRPRESSO*”, and “*mr.raps*” packages (26, 27).

## Results

As shown in Figure 2, 16,012 SNPs passed the genome-wide significance threshold (P < 5×10^-8^) in 25-Hydroxyvitamin D GWAS; after clumping those SNPs, 115 independent SNPs remained for MR analysis. Those 115 SNPs were further used in the harmonization step for hypothyroidism, Hashimoto’s thyroiditis, fT4, and TSH as the outcomes. The process of MR analyses and the results are publicly available through the following HTML link: https://akbarzadehms.github.io/VitDHypothyroidismMR/

**Figure 2.**
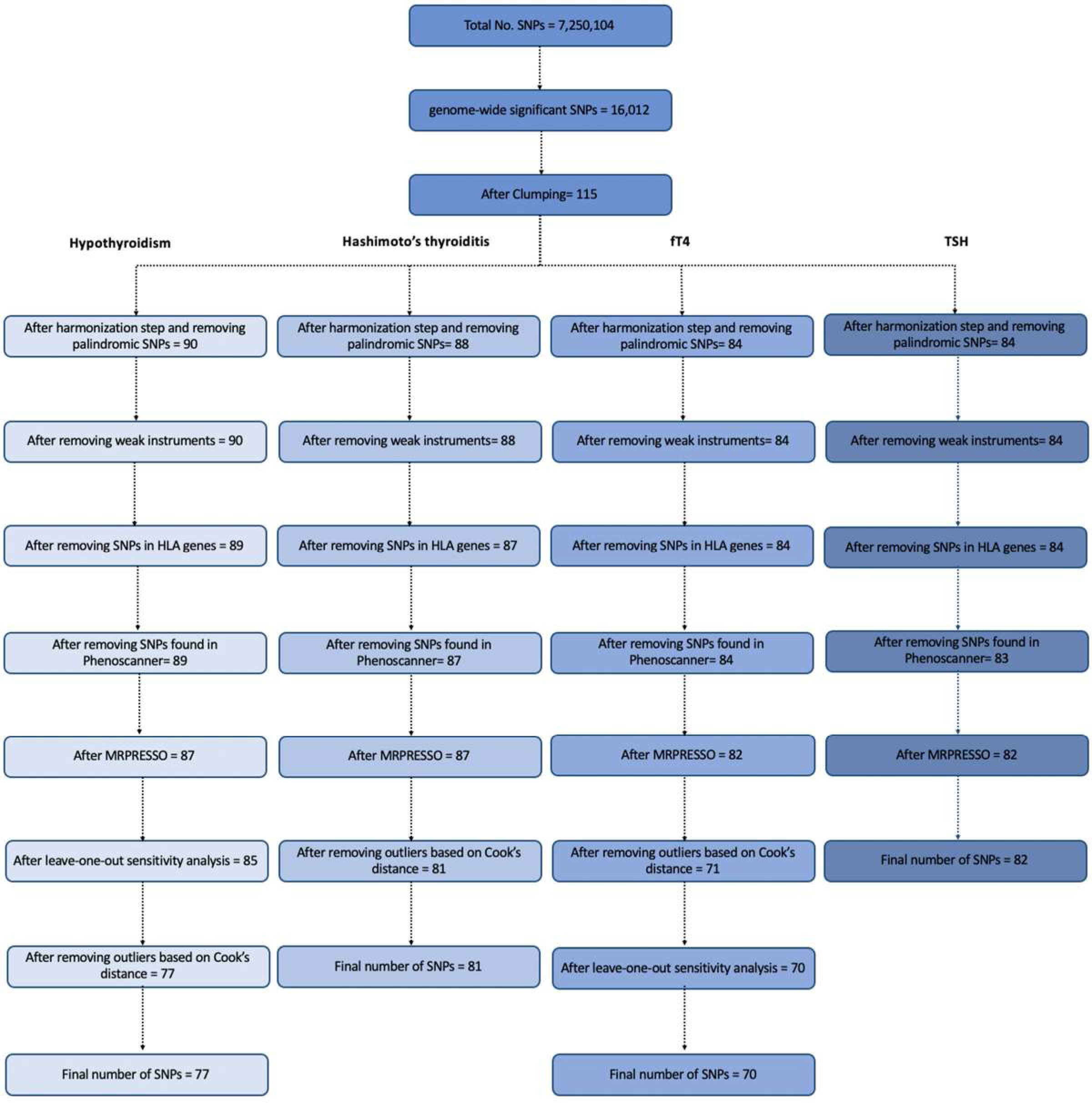
The flowchart of steps of MR analysis conducted for Hypothyroidism, Hashimoto’s thyroiditis, fT4, and TSH.

### The causal relationship between 25-Hydroxyvitamin D levels and hypothyroidism

After the harmonization step and removing palindromic SNPs, 90 SNPs were extracted. None of the remained SNPs were detected as weak instruments (F>10). One SNP was found to be in HLA genes and hence was excluded.

With 89 SNPs as IVs, according to the results of IVW, after adjustment, there was a nearly significant causal relationship between an increase in 25-Hydroxyvitamin D levels and a decrease in hypothyroidism risk (beta = − 0.131, 95% CI (− 0.245, − 0.017); SE = 0.058, P_beta_ = 0.023). According to the pleiotropy test, there was no pleiotropy (MR–Egger intercept p value = 0.826); however, the heterogeneity analysis found that there was heterogeneity in the analysis (the Q-p values of IVW and MR–Egger were 2.216×10^-8^ and 1.56×10^-8^, respectively). In sensitivity analysis, two, two, and eight SNPs were detected as outliers or influential observations using MR-PRESSO, leave-one-out sensitivity analysis, and Cook’s distance, respectively, and were excluded stepwise. After excluding those SNPs, 77 remained IVs for further MR analysis (Supplementary sheet 1). According to the results of IVW, there was a strongly significant causal relationship between an increase in 25-Hydroxyvitamin D levels and a decrease in hypothyroidism risk (beta = − 0.197, 95% CI (− 0.301, − 0.093); SE = 0.053, P_beta_ = 2.256×10^-4^). Neither pleiotropy (MR–Egger intercept p value = 0.229) nor heterogeneity (the Q-p values of IVW and MR–Egger were 0.499 and 0.515, respectively) were detected in the analysis. Moreover, the P-values of the weighted median (beta = − 0.214, 95% CI (− 0.377, − 0.051); P_beta_ = 9.937×10^−3^), weighted mode (beta = − 0.266, 95% CI (− 0.466, − 0.066); P_beta_ = 0.011) and MR–Egger (beta = − 0.319, 95% CI (− 0.540, − 0.098); P_beta_ = 0.006) methods were all less than 0.0125. The results of other MR methods are depicted as a forest plot in Figures 3a, 4a. Single SNP sensitivity analysis, leave-one-out sensitivity analysis, and funnel plots are available in supplementary Figures 1a, 2a, 3a.

**Figure 3.**
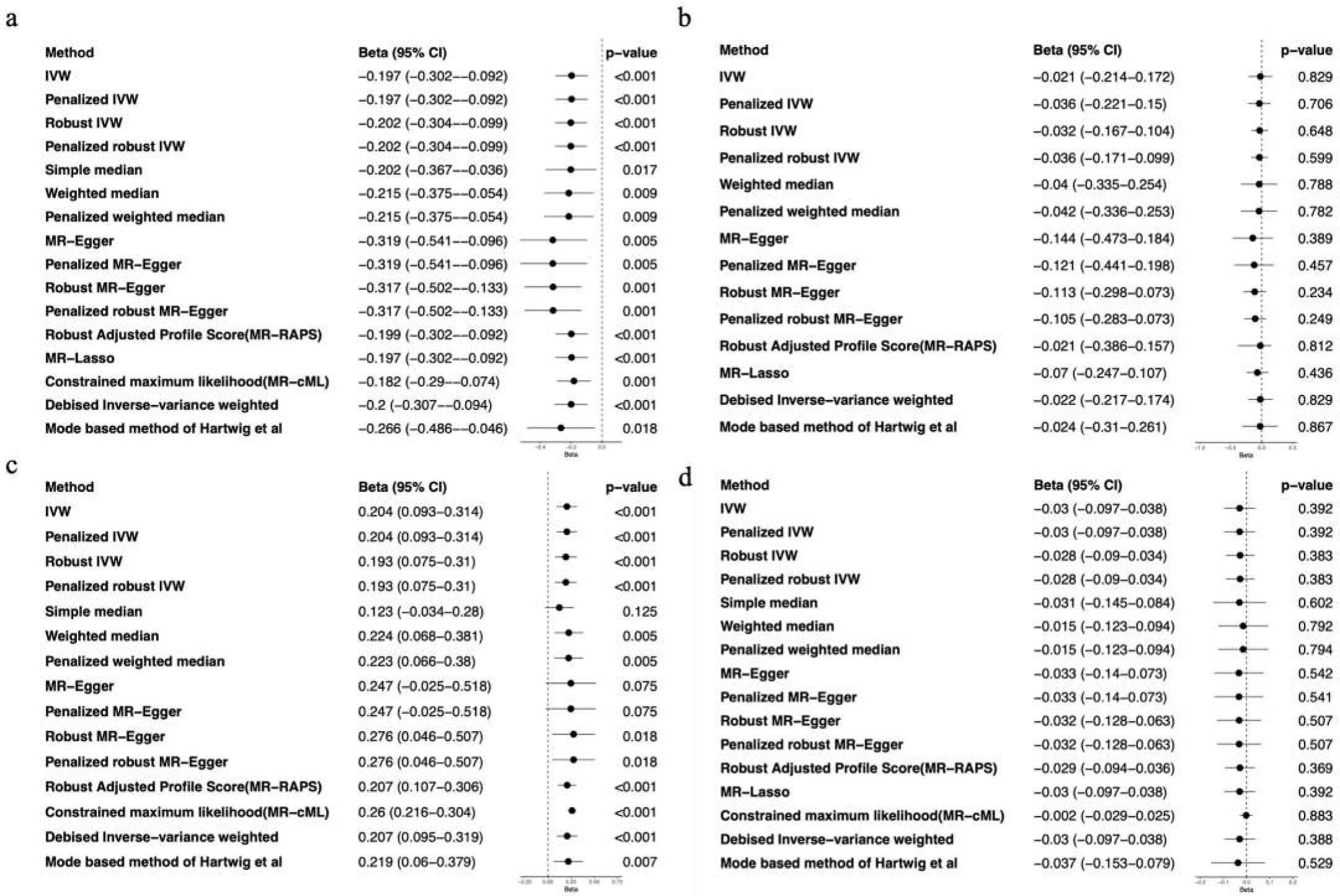
Forest plot to visualize causal effects of variation in of 25-Hydroxyvitamin D levels on a) Hypothyroidism, b) Hashimoto’s thyroiditis, c) fT4, and d) TSH. Presented odds ratios (OR) and confidence intervals (CI) correspond to the effects of 25-Hydroxyvitamin D levels on each outcome. The results of Mendelian Randomization (MR) analyses using various analysis methods are presented for comparison.

### The causal relationship between 25-Hydroxyvitamin D levels and Hashimoto’s thyroiditis

After the harmonization step and removing palindromic SNPs, 88 SNPs remained. None of the remaining SNPs were detected as weak instruments. One SNP was found to be in HLA genes and hence was excluded. With 87 SNPs as IVs, according to the results of IVW, there was no causal relationship between 25-Hydroxyvitamin D levels and Hashimoto’s thyroiditis risk (beta = - 0.047, 95% CI (− 0.245, 0.151); SE = 0.101, P_beta_ = 0.641). According to the pleiotropy test, there was no pleiotropy (MR–Egger intercept p value = 0.331); however, the heterogeneity analysis found heterogeneity in the analysis (the Q-p values of IVW and MR–Egger were 0.023 and 0.024, respectively). The sensitivity analysis excluded six SNPs detected as outliers or influential observations using Cook’s distance. In the MR analysis with 81 SNPs (Supplementary sheet 2) using various MR methods, there was no significant causal relationship between gene-predicted 25-Hydroxyvitamin D and Hashimoto’s thyroiditis after removing outlier SNPs. (The P_beta_ values in all analytical models were greater than 0.0125). Moreover, pleiotropy and heterogeneity analysis revealed that the results are not affected by pleiotropy (MR–Egger intercept p value = 0.366) or heterogeneity (the Q-p values of IVW and MR–Egger were 0.103 and 0.102, respectively). The results of all MR methods are depicted as a forest plot in Figure 3b,4b. Single SNP sensitivity analysis, leave-one-out sensitivity analysis, and funnel plots are available in supplementary Figures 1b, 2b, 3b.

**Figure 4.**
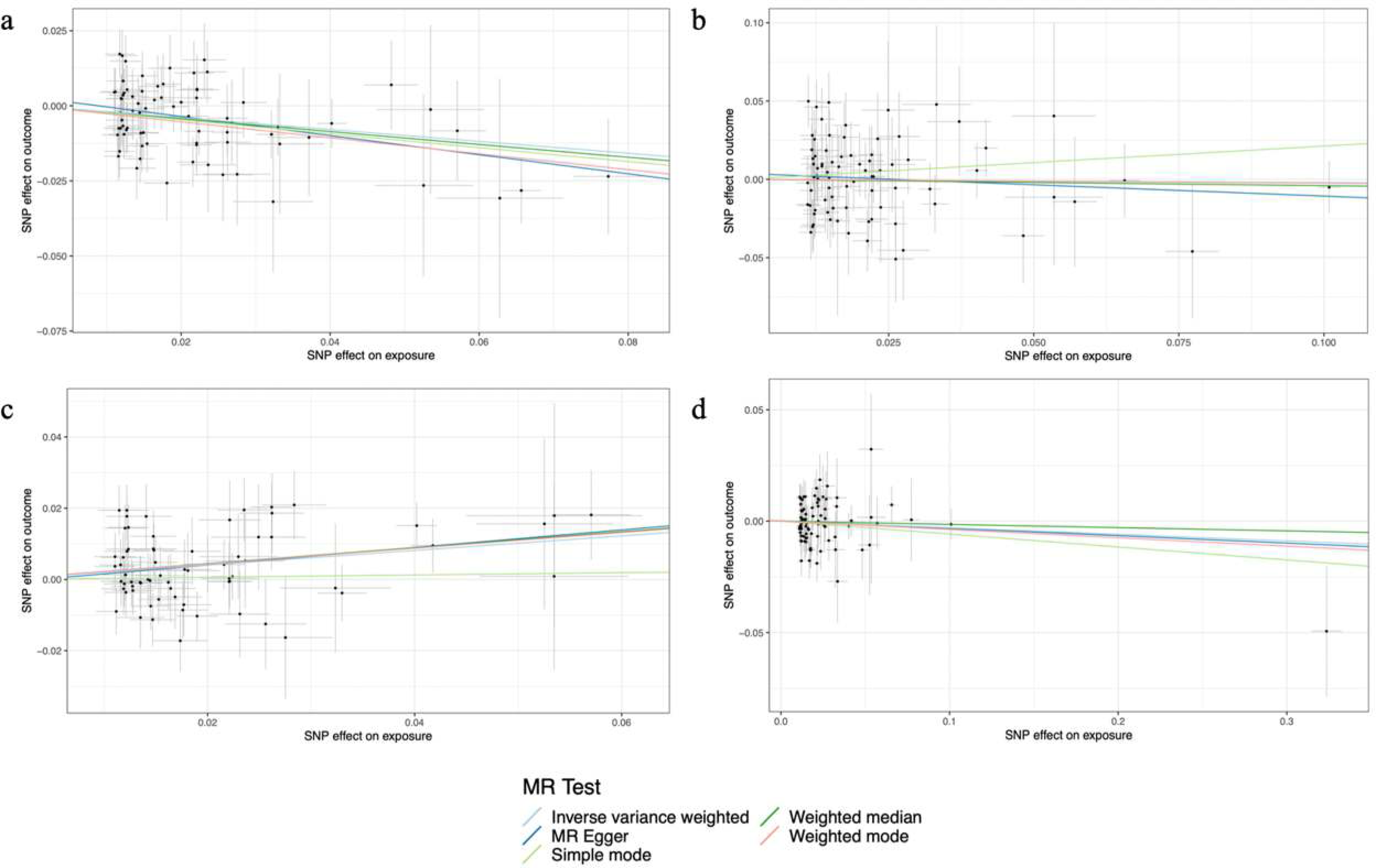
Scatter plots of 25-Hydroxyvitamin D levels on a) Hypothyroidism, b) Hashimoto’s thyroiditis, c) fT4, d) TSH. Scatter plot demonstrating the effect of each 25-Hydroxyvitamin D-associated SNP on each outcome on the log-odds scale. The slopes of each line represent the causal association for each MR method (MR-Egger, Weighted median, Inverse variance weighted, Simple mode, and Weighted mode).

### The causal relationship between 25-Hydroxyvitamin D levels and fT4

After the harmonization step and removing palindromic SNPs, 84 SNPs remained. None of the remaining SNPs were detected as weak instruments, and no SNPs were found in HLA genes. With 84 SNPs as IVs, according to the results of IVW, there was no causal relationship between 25- Hydroxyvitamin D levels and fT4 levels (beta = 0.012, 95% CI (− 0.094, 0.118); SE = 0.054, P_beta_ = 0.821). According to the pleiotropy test, there was no pleiotropy (MR–Egger intercept p value = 0.414); however, the heterogeneity analysis found heterogeneity in the analysis (the Q-p values of IVW and MR–Egger were 1.156 ×10-11 and 1.199×10-11, respectively). To correct the heterogeneity, sensitivity analyses were performed stepwise using MR-PRESSO, Cook’s distance, and leave-one-out sensitivity analysis, and two, eleven, and one SNPS were excluded in each step, respectively. After excluding the aforementioned SNPs, 70 SNPs remained and were used as IVs for further MR analysis (Supplementary sheet 3). According to the results of IVW, there was a significant causal relationship between an increase in 25-Hydroxyvitamin D levels and an increase in fT4 levels (beta = 0.204, 95% CI (0.305, 0.094); SE = 0.056, P_beta_ = 3.0506e−4). Neither pleiotropy (MR–Egger intercept p value = 0.735) nor heterogeneity (the Q-p values of IVW and MR–Egger were 0.073 and 0.064, respectively) were detected in the analysis. Moreover, the P values of the weighted median (beta = 0.223, 95% CI (0.067, 0.379); P_beta_ = 0.005) method was less than 0.0125. The results of other MR methods are depicted as a forest plot in Figures 3c, 4c. Single SNP sensitivity analysis, leave-one-out sensitivity analysis, and funnel plots are available in supplementary Figures 1c, 2c, 3c.

### The causal relationship between 25-Hydroxyvitamin D levels and TSH

After the harmonization step and removing palindromic SNPs, 84 SNPs remained. None of the remaining SNPs were detected as weak instruments, and no SNPs were found in HLA genes. One variant was found to be pleiotropic in the Phenoscanner Database and hence was excluded. One SNP was detected as an outlier in sensitivity analyses using MR-PRESSO. After excluding the mentioned SNP, 82 SNPs were used as IVs for MR analysis (Supplementary sheet 4). In the MR analysis using various MR methods, there was no significant causal relationship between gene-predicted 25-Hydroxyvitamin D and TSH (the P_beta_ values in all analytical models were greater than 0.0125).

Moreover, pleiotropy and heterogeneity analysis revealed that the results are not affected by pleiotropy (MR–Egger intercept p value = 0.931) or heterogeneity (the Q-p values of IVW and MR–Egger were 0.260 and 0.236, respectively). The results of all MR methods are depicted as a forest plot in Figures 3d and 4d. Single SNP sensitivity analysis, leave-one-out sensitivity analysis, and funnel plots are available in supplementary Figures 1d, 2d, 3d.

## Discussion

To our knowledge, this is the first study to determine the causal relationship between 25- Hydroxyvitamin D and hypothyroidism using a two-sample MR analysis. Using two-sample MR, we concluded that genetically predicted lower levels of 25-Hydroxyvitamin D have a significant causal association with the risk of hypothyroidism. In addition, the selected instrumental variables also determined a causal association between lower 25-Hydroxyvitamin D levels and lower levels of fT4 hormone. There is a relatively high clinical implication for this causality due to the high worldwide prevalence of 25-Hydroxyvitamin D deficiency and the ease of its control using oral supplements in most cases (28).

The association between 25-Hydroxyvitamin D levels and hypothyroidism has been investigated in several studies. Mackawy et al. conducted an observational study on 30 cases of hypothyroidism, diagnosed according to biochemical criteria, and 30 healthy controls from the Saudi Arabian population. The serum levels of 25-Hydroxyvitamin D were measured in both cases and controls. The results showed significantly lower levels of 25-Hydroxyvitamin D in hypothyroidism cases compared to the control group (t=-11.128, p<0.001). However, the subgroup analysis did not reveal any significant differences between male and female subjects (t=-1.32, p>0.05) (29). In a study on summertime 25-Hydroxyvitamin D status in patients with thyroid disease in Poland, results indicated significantly lower levels of the 25-Hydroxyvitamin D metabolites 25(OH)D and 3-epi-25(OH)D in hypothyroid patients in comparison with euthyroid cases (30). Higher than 125 nmol/L serum levels of 25(OH)D in a secondary analysis of data of the Pure North program in Canada is associated with a lower risk of hypothyroidism and a reduction in levels of anti-thyroid antibodies by 30% and 32%, respectively, according to Mirhosseini et al. (31). Although, a study by Musa et al. on a Saudi population showed no significantly different levels of 25-Hydroxyvitamin D in female patients with a hypothyroidism diagnosis in comparison with healthy controls (32). Suggesting the differences in results depending on the population subgroup investigated in the study.

Autoimmune thyroid diseases (AITD) are the most common form of organ-specific autoimmune diseases with an estimated prevalence of 5%, including a vast spectrum of manifestations ranging from hyperthyroidism, known as Graves’ disease, to hypothyroidism, known as Hashimoto’s thyroiditis, as well as cases with no history of clinical and biochemical findings (33, 34). The pathophysiology of AITDs has been described as dysfunction of the normally established tolerance towards thyroid tissue antigens, infiltration of lymphocytes, subsequent activation of the inflammatory response, and production of the characteristic autoantibodies. The commonly targeted autoantigens are thyroglobulin (Tg) and thyroid peroxidase (TPO) in Hashimoto’s thyroiditis and Thyroid stimulating hormone (TSH) receptor (TSHR) in Graves’ disease (34, 35). In addition to the studies on the association of 25-Hydroxyvitamin D and hypothyroidism of various etiologies, several studies have investigated this association in Hashimoto’s thyroiditis patients. *In vivo,* studies have shown that treatment of rat models of experimental autoimmune thyroiditis (EAT) with 1,25(OH)_2_D3 prior to disease establishment is associated with maintaining normal structural and biochemical thyroid profiles (36). The results of clinical observational studies on this matter have shown substantial controversy, with both indicating insignificant association as well as a significant inverse correlation between 25-Hydroxyvitamin D levels and the risk of Hashimoto’s thyroiditis (17, 37–39). A recent meta-analysis of 42 observational studies also indicated significantly lower levels of serum 25-Hydroxyvitamin D in patients of hypothyroidism (p=0.03), AITDs (p=0.013), and Hashimoto’s thyroiditis (p<0.001) in comparison with controls (40).

In addition to the aforementioned observational studies, few clinical trials have been conducted on this matter. A randomized controlled trial (RCT) was conducted by Chahardoli et al. on 42 female cases of Hashimoto’s thyroiditis from an Iranian population. The patients were divided into intervention, receiving weekly 25-Hydroxyvitamin D supplements, and control groups, receiving placebo. The results of this study indicated a significant reduction of levels of TSH and anti-Tg antibody in the treatment group over the course of the study. No significant changes were observed in levels of anti-TPO, T3, and T4 (18). In a pilot randomized trial by Pezeshki et al., the effects of 25-Hydroxyvitamin D supplements were evaluated in 59 cases with the coexistence of subclinical hypothyroidism and 25-Hydroxyvitamin D deficiency in an Iranian population. Also, they showed a reduction in TSH levels throughout the study (41). In another RCT in an Iranian population on 201 cases of hypothyroidism, 25-Hydroxyvitamin D supplements significantly reduced TSH levels in the treatment group with no significant alterations in levels of T3 and T4 (14). Although the effect of 25-Hydroxyvitamin D supplements on thyroid function in hypothyroidism patients has been investigated in the studies mentioned above, there is substantial controversy among the results. To the best of our knowledge, the association of 25-Hydroxyvitamin D with the risk of hypothyroidism has not been investigated in RCTs, and due to the limitations of such studies, MR studies can be an appropriate substitute to establish this causality. A recently published bidirectional MR study investigated the causal relationship between levels of 25-Hydroxyvitamin D and anti-TPO antibody using data from a Chinese population. The results of that study indicated a significant causal relationship (*β*= −0.720, 95% CI −1.429 to −0.012) between levels of 25- Hydroxyvitamin D and anti-TPO antibody (42).

Our results indicated a causal relationship between lower 25-Hydroxyvitamin D levels and both the risk of hypothyroidism and lower levels of fT4, which is consistent with the results of the aforementioned observational studies. However, we found no significant causal association between this parameter and TSH levels or risk of Hashimoto’s thyroiditis. As mentioned above, the results of the previously conducted studies are controversial and differ based on the target population, number of included patients, and study design. Although the previously mentioned clinical trials found significant associations between vitamin D and TSH levels, the results are not necessarily accurate due to the limited sample sizes and non-European ancestry of the included patients, in contrast with this MR study.

It is known that the most common cause of hypothyroidism in the world and the European population is iodine deficiency. In iodine-sufficient populations, Hashimoto’s thyroiditis is the most common cause of hypothyroidism (7). Since in our study, there was no significant relationship between 25-Hydroxyvitamin D deficiency and Hashimoto’s thyroiditis, it can be speculated that 25-Hydroxyvitamin D deficiency has a causal relationship with other hypothyroidisms and even non-autoimmune etiologies and since the iodine deficiency is the leading cause of hypothyroidism, future MR studies should be conducted to infer the causal relationship of 25-Hydroxyvitamin D with iodine deficiency.

Several mechanisms have been proposed in the explanation of such associations. Both 25- Hydroxyvitamin D and thyroid hormones carry out their functional roles by binding to members of the steroid/thyroid receptor family (43). In addition, 25-Hydroxyvitamin D is a modulator of TSH secretion in the pituitary gland, both directly by binding to specific binding sites and utilizing the regulation of estrogen synthesis, which is a modulator of TSH secretion (44, 45). Regarding its role in AITDs’ pathogenesis, 25-Hydroxyvitamin D is a modulator of the immune system. Both VDR and 1-a hydroxylase are expressed by a variety of immune cells including the antigen-presenting cells (APCs) and lymphocytes (46). Activation of 25-Hydroxyvitamin D downstream effectors leads to inhibition of expression of the major histocompatibility complex II (MHC II) and its co-stimulators in APCs, and interleukin-12 (IL-12) and IL-23 in dendritic cells and induction of IL-10 expression, ultimately leading to a reduction in antigen presentation and T cell activation, as well as a shift in T cell phenotypes towards T helper 2 (Th2) cells (47, 48). In B cells, 25-Hydroxyvitamin D inhibits the proliferation, memory B, and plasma cell differentiation, and immunoglobulin secretion and induces apoptotic processes. In addition to the aforementioned roles in inhibiting the adaptive immune response, 25-Hydroxyvitamin D is an inducer of innate immune reactions (46).

Regarding clinical relevance, our study established a causal relationship between lower levels of 25-Hydroxyvitamin D and hypothyroidism, suggesting a protective effect of 25-Hydroxyvitamin D supplementation in reducing the risk of hypothyroidism in the population. This can call for public policy decisions in this matter due to the high effect size and low cost of such population-wide plans to initiate screening and treating 25-Hydroxyvitamin D deficiency.

This study possesses several strengths. First, the data regarding the instrumental variables are from reliable and widely used GWAS databases with acceptably large sample sizes. Second, the two-sample MR is a method of analysis that substantially reduces the effect of confounders by using genetic variants as instrumental variables. Finally, the sensitivity analyses and estimations of pleiotropy reduce the risk of bias due to the presence of outliers and pleiotropic effects of the instrumental variables.

Despite the strengths above, this study faced some limitations as well. The data regarding the instrumental variables are extracted from European population GWASs, which can limit the application of our results in other populations, and the results should be interpreted cautiously. Second, the GWASs primarily used to extract the summary statistic data might impose some limitations on our study, including population stratification and relatedness. Although measures were taken to reduce and omit such limitations, there is a possibility that these sources of bias still exist. Overall, although our results are generally applicable in various conditions, the results should be interpreted with caution.

## Conclusion

In summary, this study provides evidence for a possible causal association between lower levels of vitamin D and a higher risk of hypothyroidism and lower free T4 levels, using genetic variations as an instrumental variable. These findings can be used in future population-level policies and plans to prevent and manage hypothyroidism.

## Availability of data and materials

The datasets analyzed during the current study are publicly available and were extracted from the GWAS catalog database and the ThyroidOmics Consortium. The process of MR analyses and the results are publicly available through the following HTML link: https://akbarzadehms.github.io/VitDHypothyroidismMR/

## Abbreviations

CI: Confidence interval
GWAS: Genome-Wide Association Study
IVs: Instrumental Variables
IVW: Inverse Variance-Weighted
MAF: Mean Allele Frequency
MBE: Mode-Based Estimation
MR: Mendelian Randomization
MR-PRESSO: Mendelian Randomization Pleiotropy RESidual Sum and Outlier
RCT: Randomized Clinical Trial
SE: Standard Error
SNP: Single Nucleotide Polymorphism
TSH: Thyroid Stimulating Hormone
fT4: free T4
VDR: 25-Hydroxyvitamin D Receptor

## Contributions

MA and STF conceptualize the study. MA, AM, and STF drafted the initial manuscript. MA and STF conceived the study, analyzed the data, and drafted the initial manuscript. MA and STF generated Tables and Figures. DH, AHG, AHS, validation, and revised the manuscript. PR, HL, MJ, FT, FA, MH, and MMJ revised the manuscript. MSD supervised, edited, and finalized the manuscript. All authors reviewed and approved the final manuscript.

## Ethics declarations

The ethical committee approved this study at the Research Institute for Endocrine Sciences, Shahid Beheshti University of Medical Sciences (Research Approval Code: 28778 & Research Ethical Code: IR.SBMU.ENDOCRINE.REC.1400.084). In this study, all participants provided written informed consent for participating in the study. This study has been performed following the Declaration of Helsinki.

## Consent for publication

Not applicable.

