## Supplementary material for "Genetically predicted 25-Hydroxyvitamin D levels on Hypothyroidism: A two-sample Mendelian Randomization": supplemantry figures

Running title: **Causal Relationship between 25-Hydroxyvitamin D Levels and Hypothyroidism**


**Authors:**

1. Mahdi Akbarzadeh; Ph.D., Cellular and Molecular Endocrine Research Center, Research Institute for Endocrine Sciences, Shahid Beheshti University of Medical Sciences, Tehran, Iran.
2. Sahand Tehrani Fateh; MD, School of Medicine, Tehran University of Medical Sciences, Tehran, Iran.
3. Aysan Moeinafshar, MD, School of Medicine, Tehran University of Medical Sciences, Tehran, Iran.
4. Danial Habibi; Ph.D., Cellular and Molecular Endocrine Research Center, Research Institute for Endocrine Sciences, Shahid Beheshti University of Medical Sciences, Tehran, Iran.
5. Amir Hossein Ghanooni; MD, Department of Endocrinology, School of Medicine, Iran University of Medical Sciences, Tehran, Iran.
6. Hesam Saeidian; MD, Department of Surgery, Rasool-E Akram Hospital, School of Medicine, Iran University of Medical Sciences, Tehran, Iran.
7. Parisa Riahi; MSc, Cellular and Molecular Endocrine Research Center, Research Institute for Endocrine Sciences, Shahid Beheshti University of Medical Sciences, Tehran, Iran.
8. Maryam Zarkesh: Cellular and Molecular Endocrine Research Center, Research Institute for Endocrine Sciences, Shahid Beheshti University of Medical Sciences, Tehran, Iran.
9. Hossein Lanjanian; Cellular and Molecular Endocrine Research Center, Research Institute for Endocrine Sciences, Shahid Beheshti University of Medical Sciences, Tehran, Iran.
10. Maryam Moazzam-Jazi, Cellular and Molecular Endocrine Research Center, Research Institute for Endocrine Sciences, Shahid Beheshti University of Medical Sciences, Tehran, Iran.
11. Mina Jahangiri; Ph.D., Department of Biostatistics, Faculty of Medical Sciences, Tarbiat, Modares University, Tehran, Iran.
12. Farshad Teymoori, Nutrition and Endocrine Research Center, Research Institute for Endocrine Sciences, Shahid Beheshti, Tehran, Iran.
13. Fereidoun Azizi; MD, Endocrine research center, Research Institute for Endocrine Sciences, Shahid Beheshti University of Medical Sciences, Tehran, Iran.
14. Mehdi Hedayati; Ph.D., Cellular and Molecular Endocrine Research Center, Research Institute for Endocrine Sciences, Shahid Beheshti University of Medical Sciences, Tehran, Iran.
15. Maryam Sadat Daneshpour; Ph.D., Cellular and Molecular Endocrine Research Center, Research Institute for Endocrine Sciences, Shahid Beheshti University of Medical Sciences, Tehran, Iran.

**Corresponding author:**
Maryam Sadat Daneshpour (Ph.D.), Associate Professor.
Cellular and Molecular Endocrine Research Center, Research Institute for Endocrine Sciences, Shahid Beheshti University of Medical Sciences.

**Table of content:**

Figure S1: Forest plot of variant specific inverse variance estimates for causal association between 25-Hydroxyvitamin D levels Hypothyroidism

Figure S2: Leave-one-out plot to assess if a single variant is driving the association between 25-Hydroxyvitamin D levels Hypothyroidism

Figure S3. Funnel plot of causal association between 25-Hydroxyvitamin D levels Hypothyroidism

Figure S1: Forest plot of variant specific inverse variance estimates for causal association
between 25-Hydroxyvitamin D levels Hypothyroidism

A) Hypothyroidism

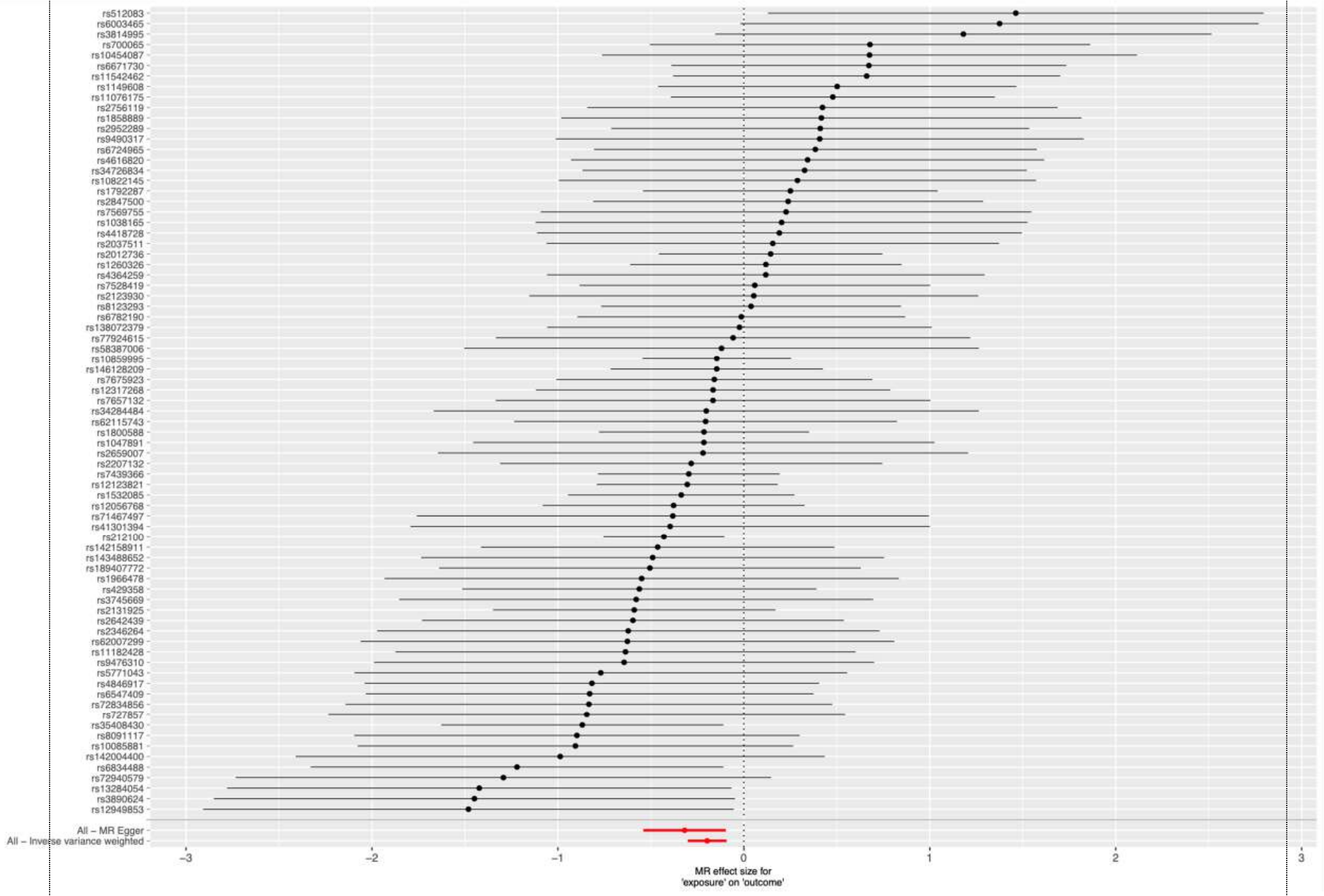

B) Hashimoto’s thyroiditis

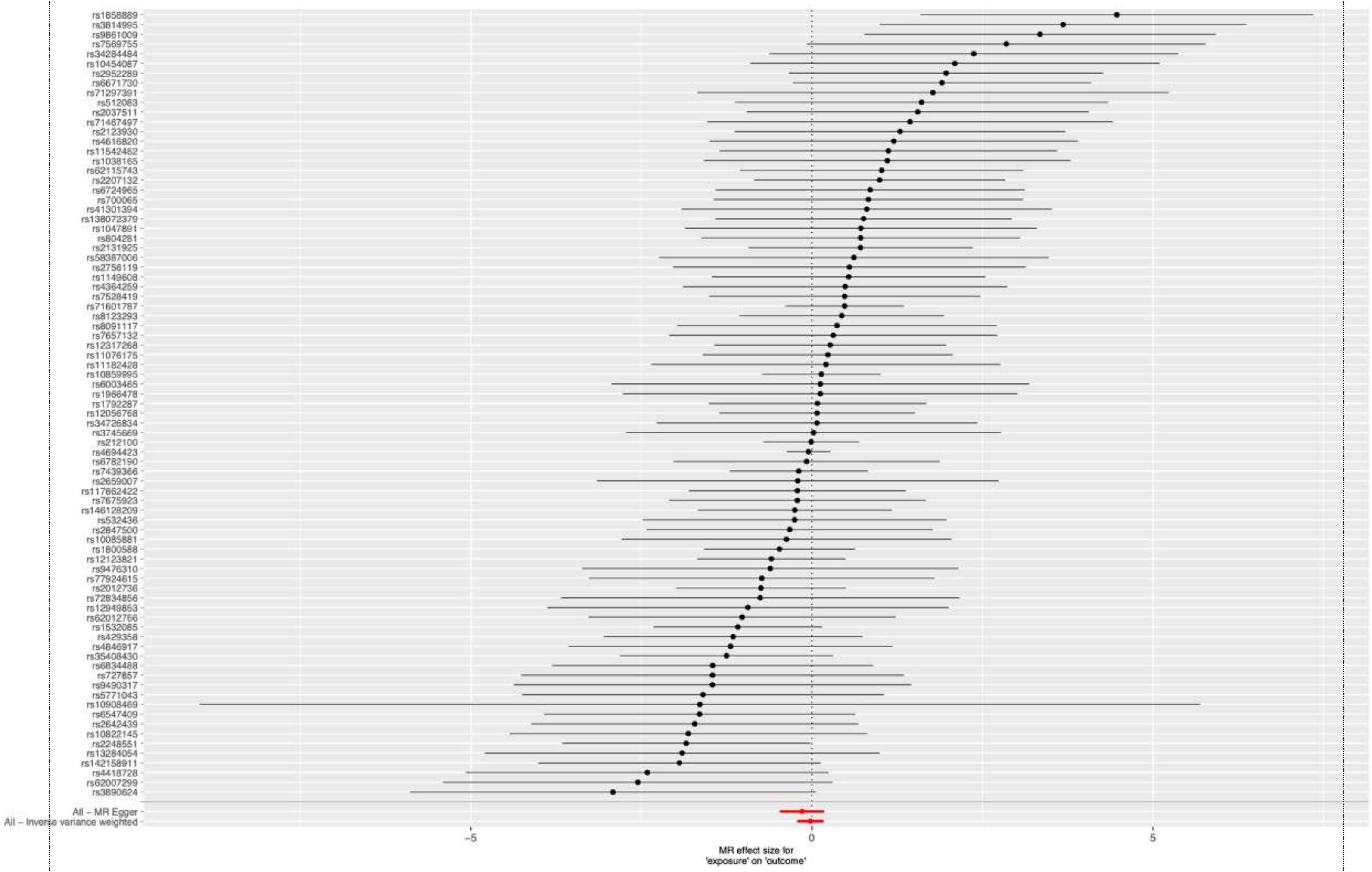


C) Free T4

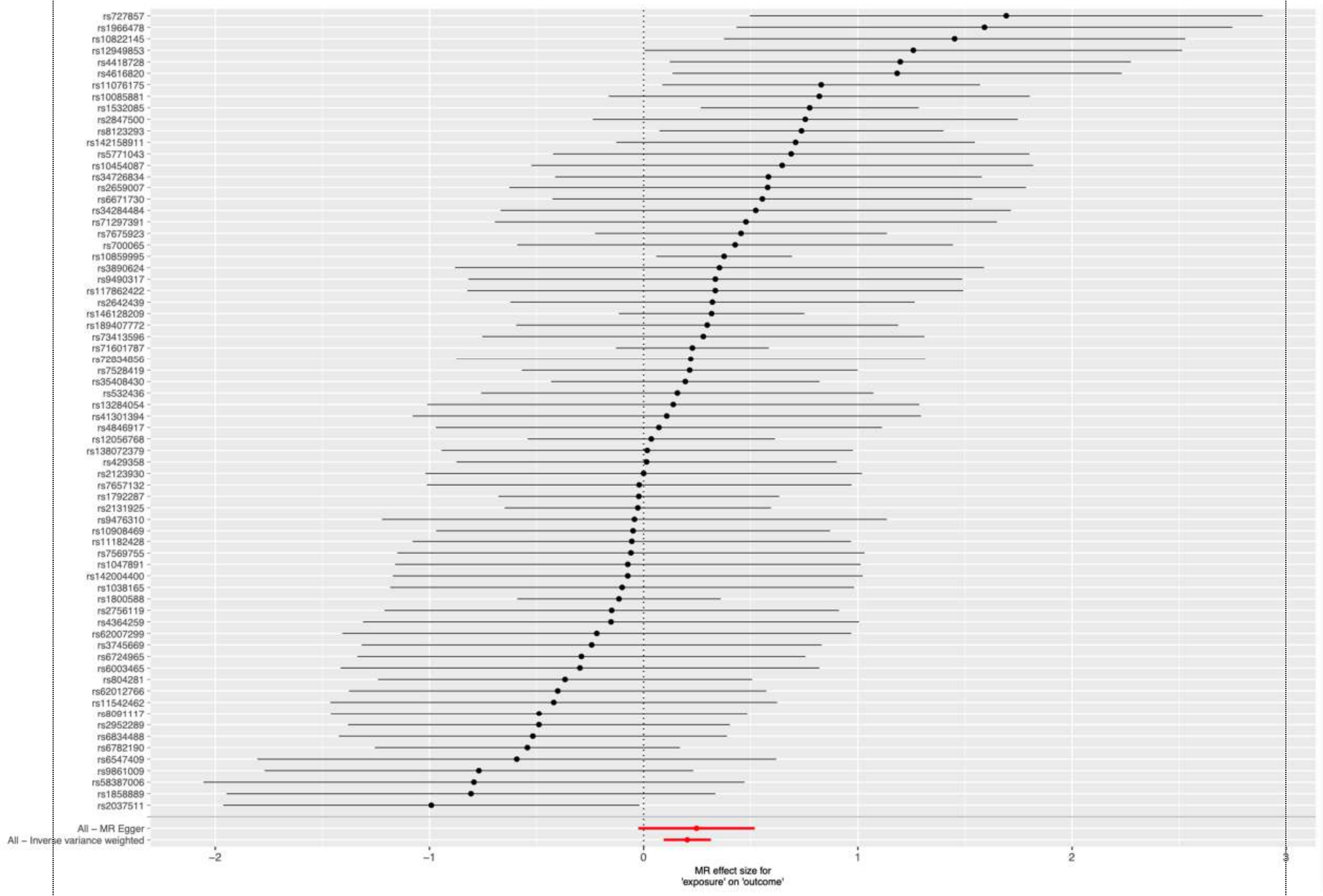

rs727857
rs1966478
rs10822145
rs12949853
rs4418728
rs4616820
rs11076175
rs10085881
rs1532085
rs2847500
rs8123293
rs142158911
rs5771043
rs10454087
rs34726834
rs2659007
rs6671730
rs34284484
rs71297391
rs7675923
rs700065
rs10859995
rs3890624
rs9490317
rs117862422
rs2642439
rs146128209
rs189407772
rs73413596
rs71601787
rs72834856
rs7528419
rs35408430
rs532436
rs13284054
rs41301394
rs4846917
rs12056768
rs138072379
rs429358
rs2123930
rs7657132
rs1792287
rs2131925
rs9476310
rs10908469
rs11182428
rs7569755
rs1047891
rs142004400
rs1038165
rs1800588
rs2756119
rs4364259
rs62007299
rs3745669
rs6724965
rs6003465
rs804281
rs62012766
rs11542462
rs8091117
rs2952289
rs6834488
rs6782190
rs6547409
rs9861009
rs58387006
rs1858889
rs2037511
All – MR Egger
All – Inverse variance weighted
−2
−1
0
1
2
3
MR effect size for
'exposure' on 'outcome'


D) TSH

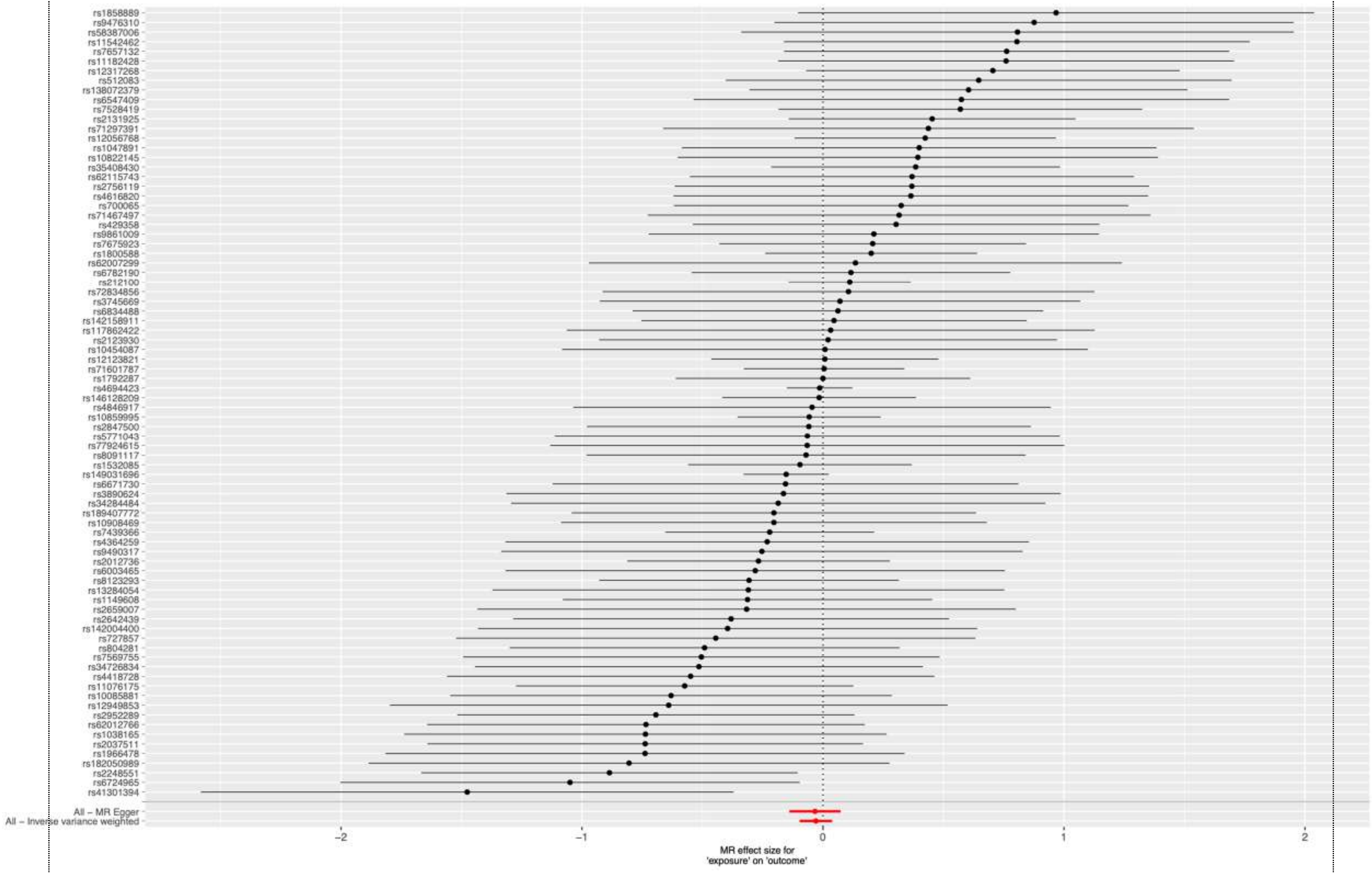
rs1858889
rs9476310
rs58387006
rs11542462
rs7657132
rs11182428
rs12317268
rs512083
rs138072379
rs6547409
rs7528419
rs2131925
rs71297391
rs12056768
rs1047891
rs10822145
rs35408430
rs62115743
rs2756119
rs4616820
rs700065
rs71467497
rs429358
rs9861009
rs7675923
rs1800588
rs62007299
rs6782190
rs212100
rs72834856
rs3745669
rs6834488
rs142158911
rs117862422
rs2123930
rs10454087
rs12123821
rs71601787
rs1792287
rs4694423
rs146128209
rs4846917
rs10859995
rs2847500
rs5771043
rs77924615
rs8091117
rs1532085
rs149031696
rs6671730
rs3890624
rs34284484
rs189407772
rs10908469
rs7439366
rs4364259
rs9490317
rs2012736
rs6003465
rs8123293
rs13284054
rs1149608
rs2659007
rs2642439
rs142004400
rs727857
rs804281
rs7569755
rs34726834
rs4418728
rs11076175
rs10085881
rs12949853
rs2952289
rs62012766
rs1038165
rs2037511
rs1966478
rs182050989
rs2248551
rs6724965
rs41301394
All – MR Egger
All – Inverse variance weighted
−2
−1
0
1
2
MR effect size for
'exposure' on 'outcome'

**Figure S2:** Leave-one-out plot to assess if a single variant is driving the association between 25-Hydroxyvitamin D levels Hypothyroidism

A) Hypothyroidism loo

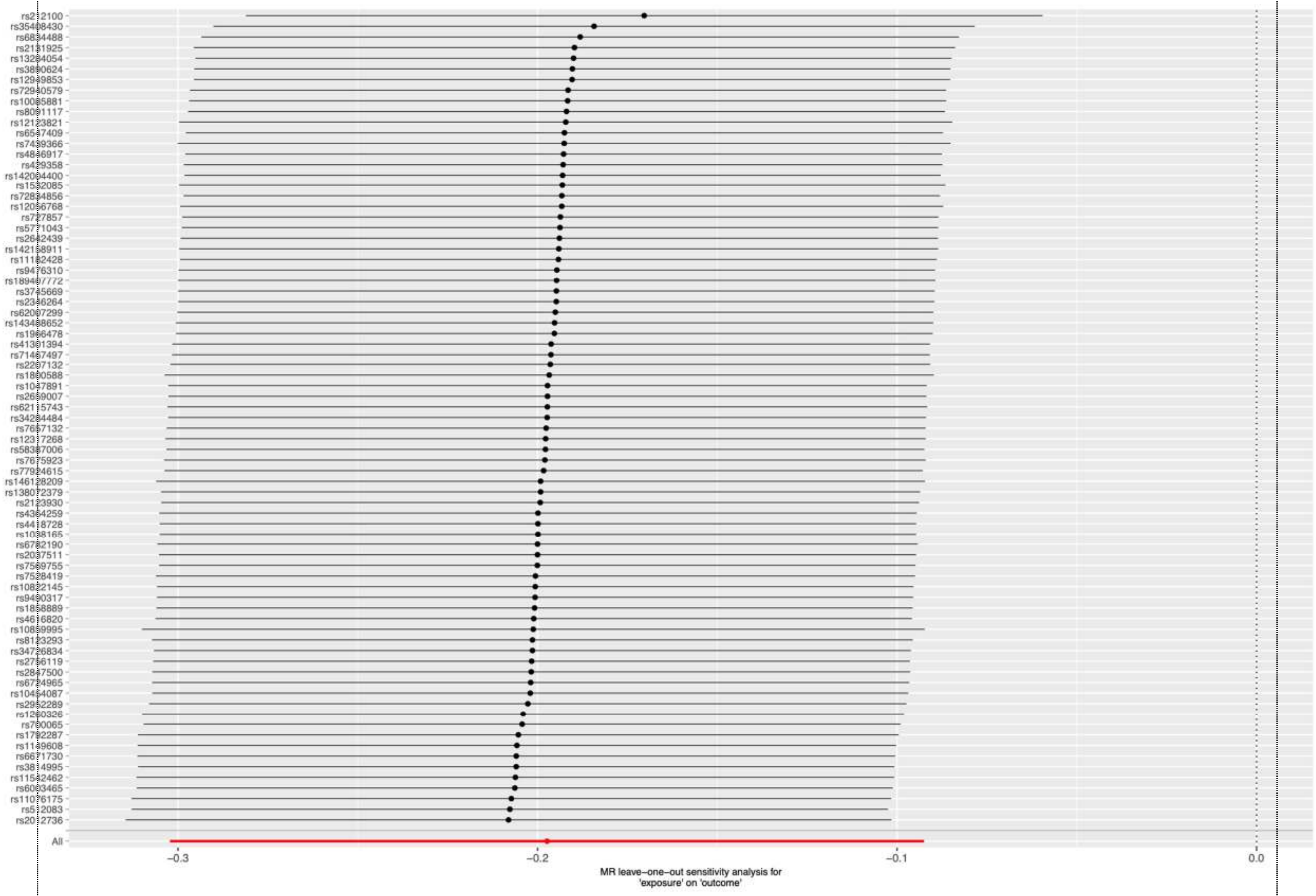


B) Hashimoto's thyroiditis

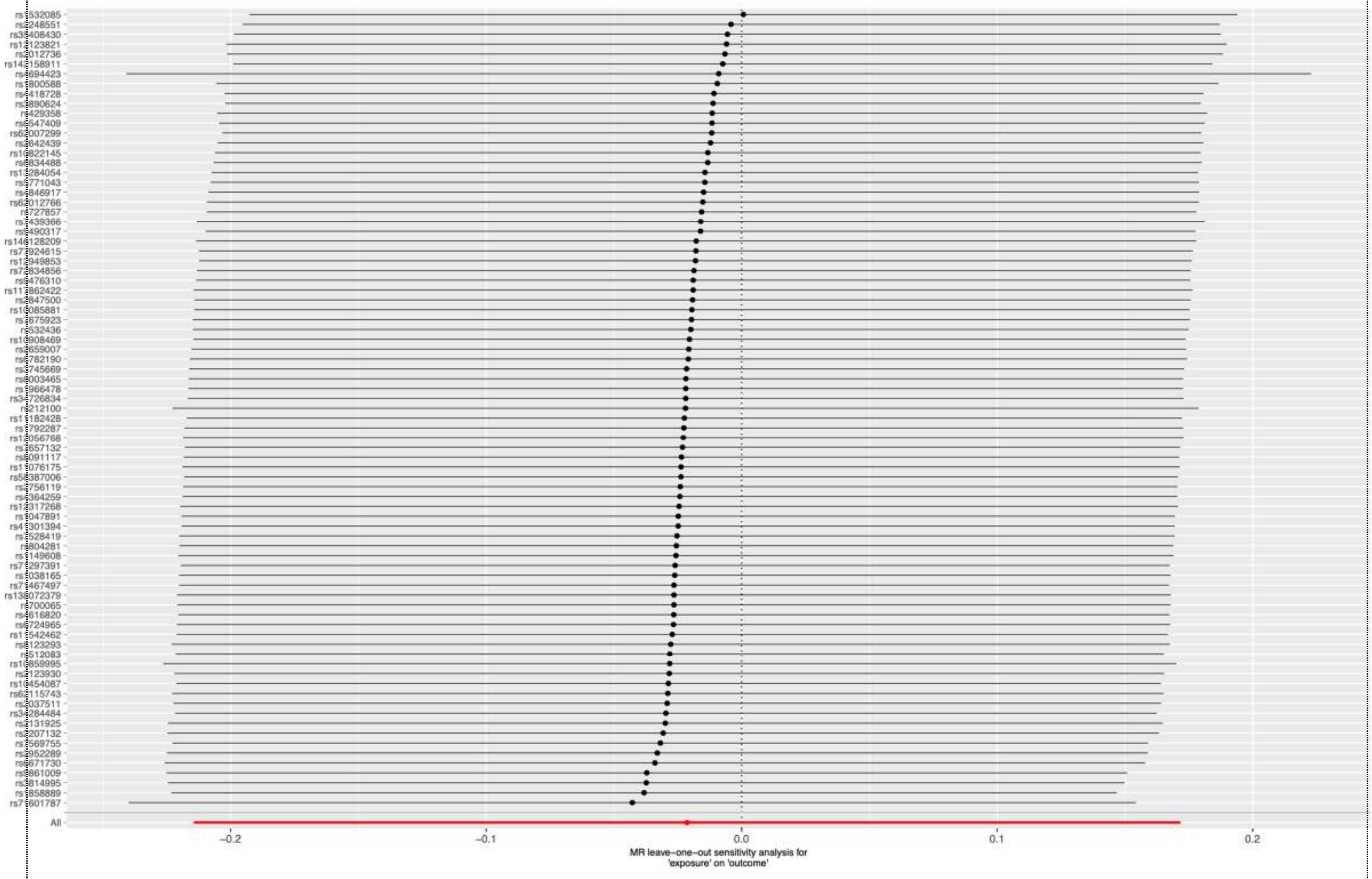
All
−0.2
−0.1
0.0
0.1
0.2
MR leave−one−out sensitivity analysis for
'exposure' on 'outcome'

C) Free T4

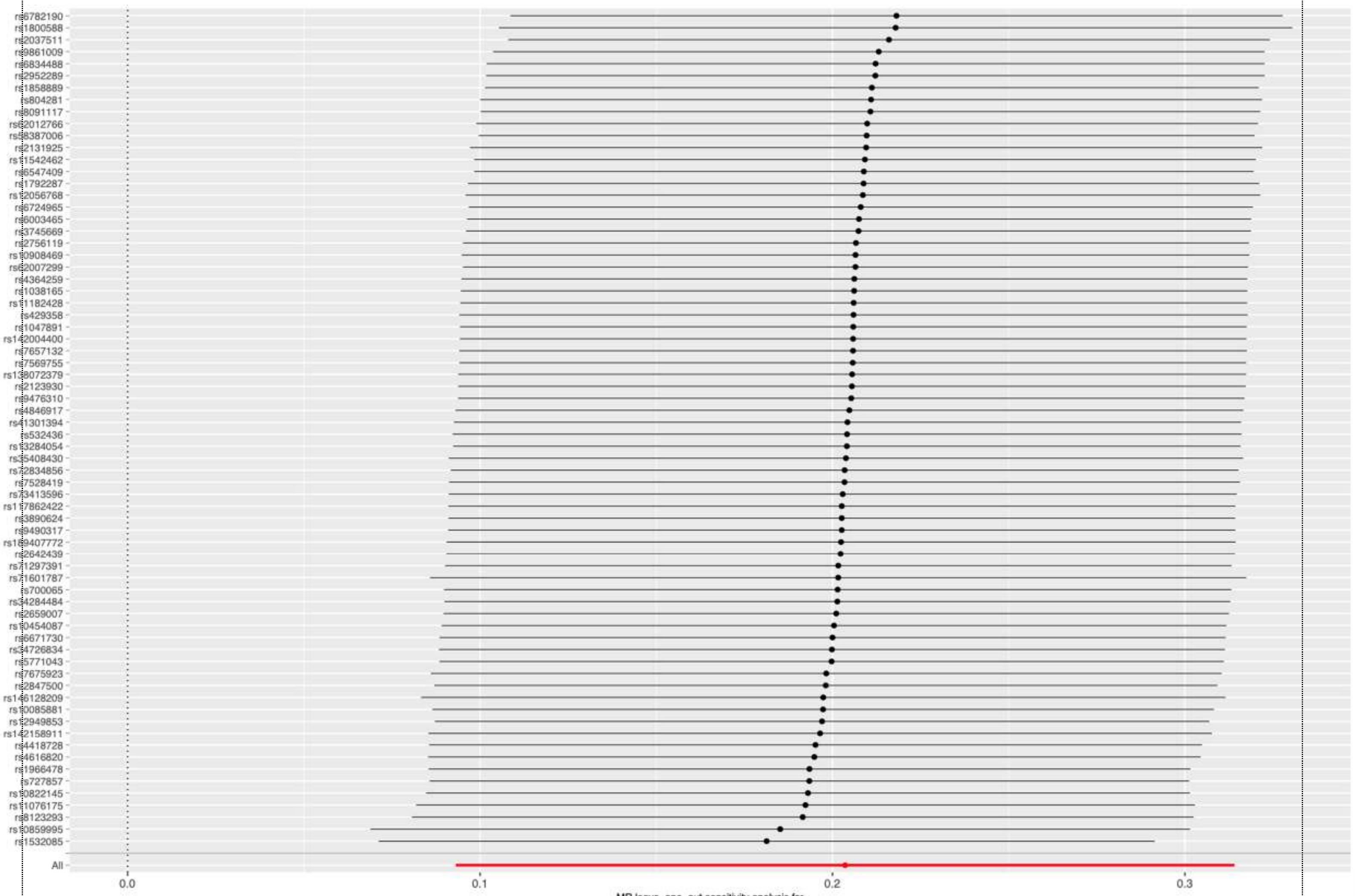


MR leave-one-out sensitivity analysis for
'exposure' on 'outcome'

D) TSH

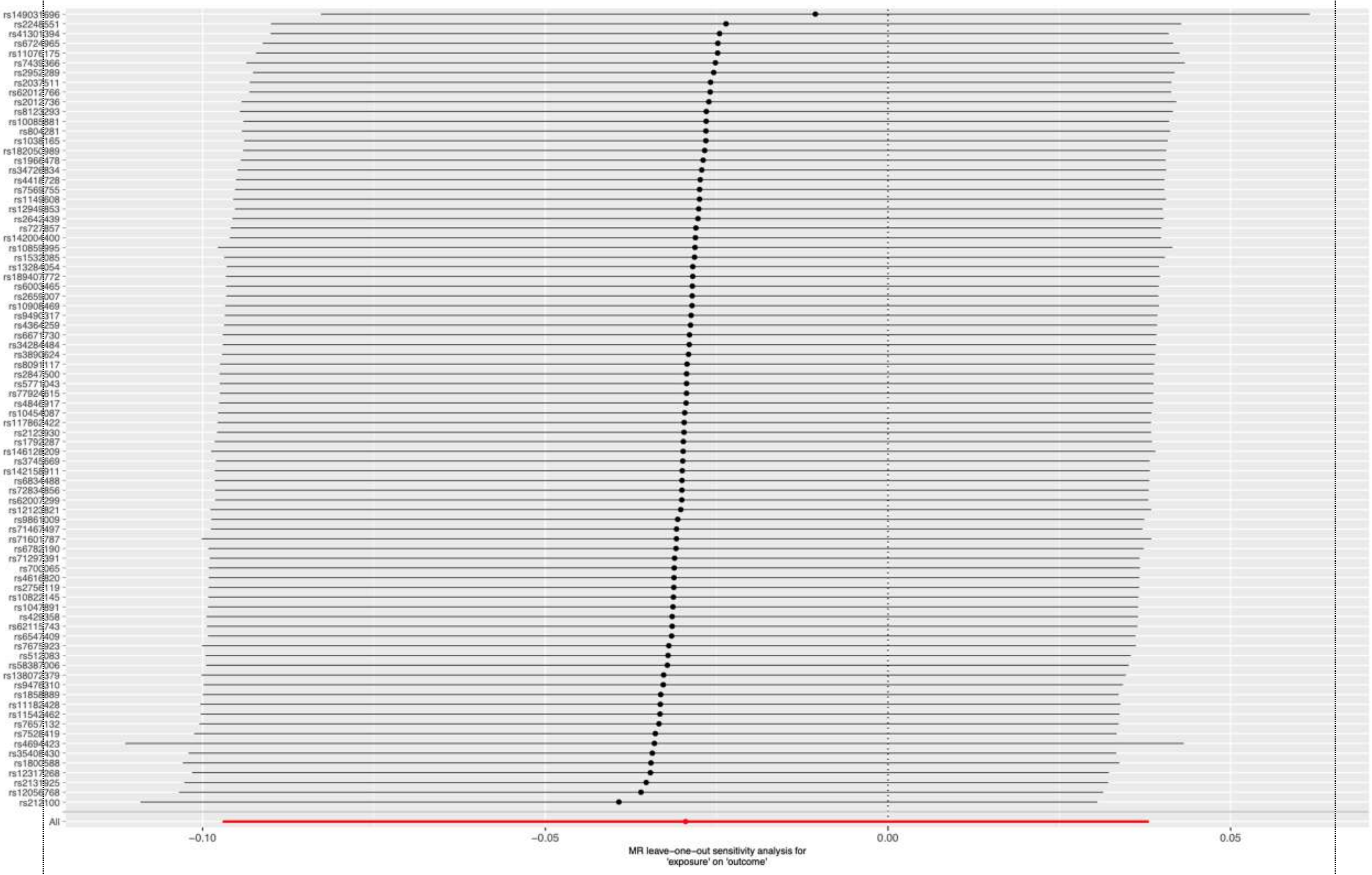


**Figure S3.** Funnel plot of causal association between 25-Hydroxyvitamin D levels Hypothyroidism

A) Hypothyroidism

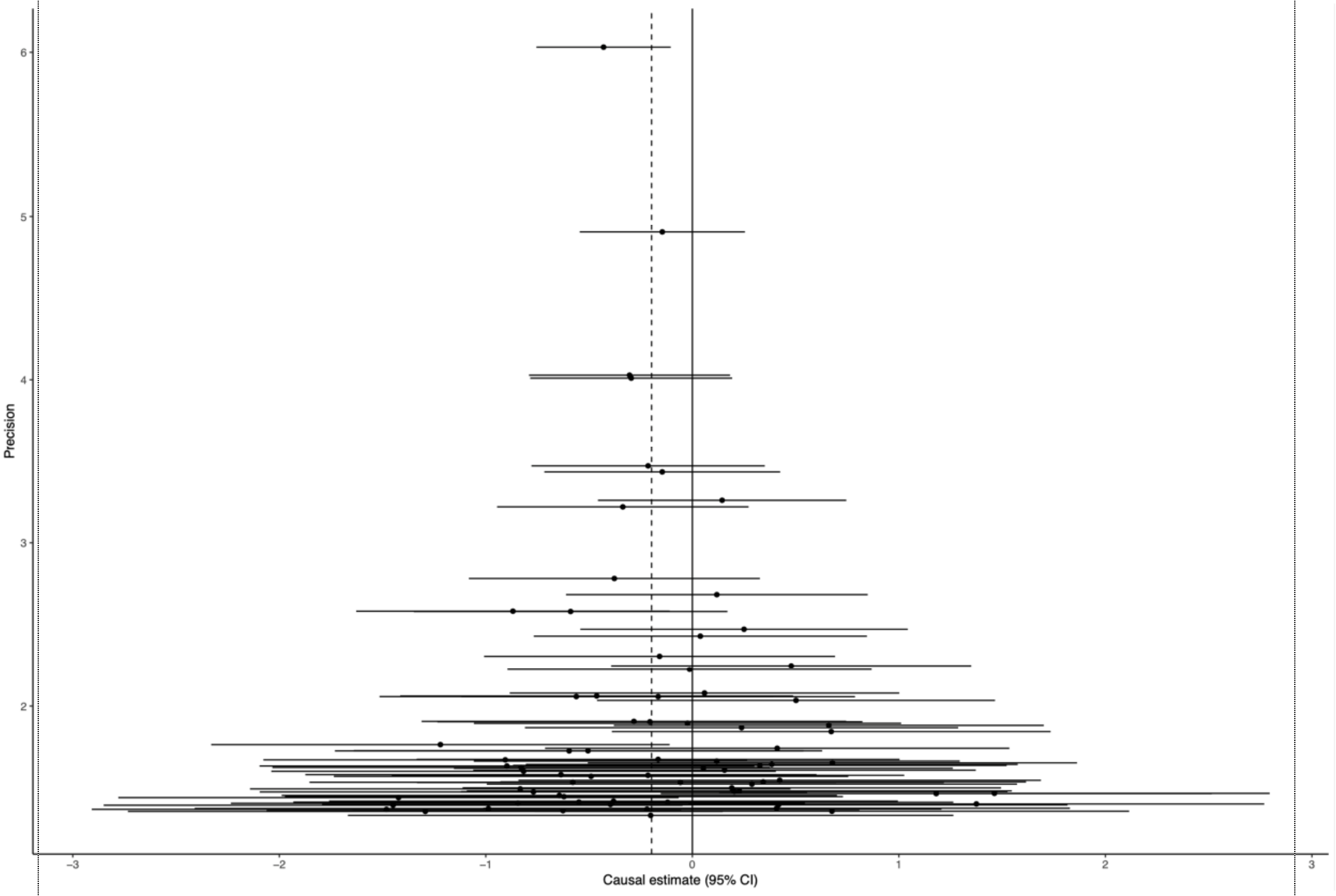


B) Hashimoto’s thyroiditis

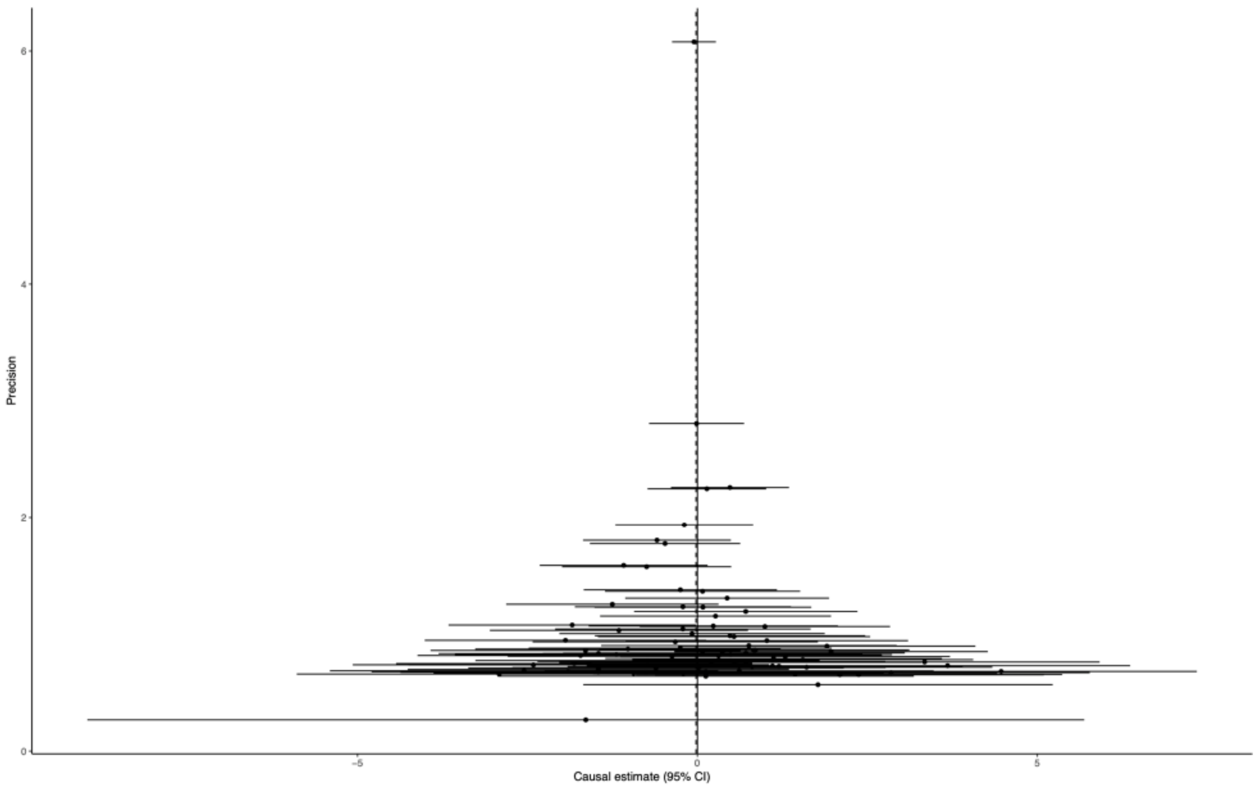


C) Free T4

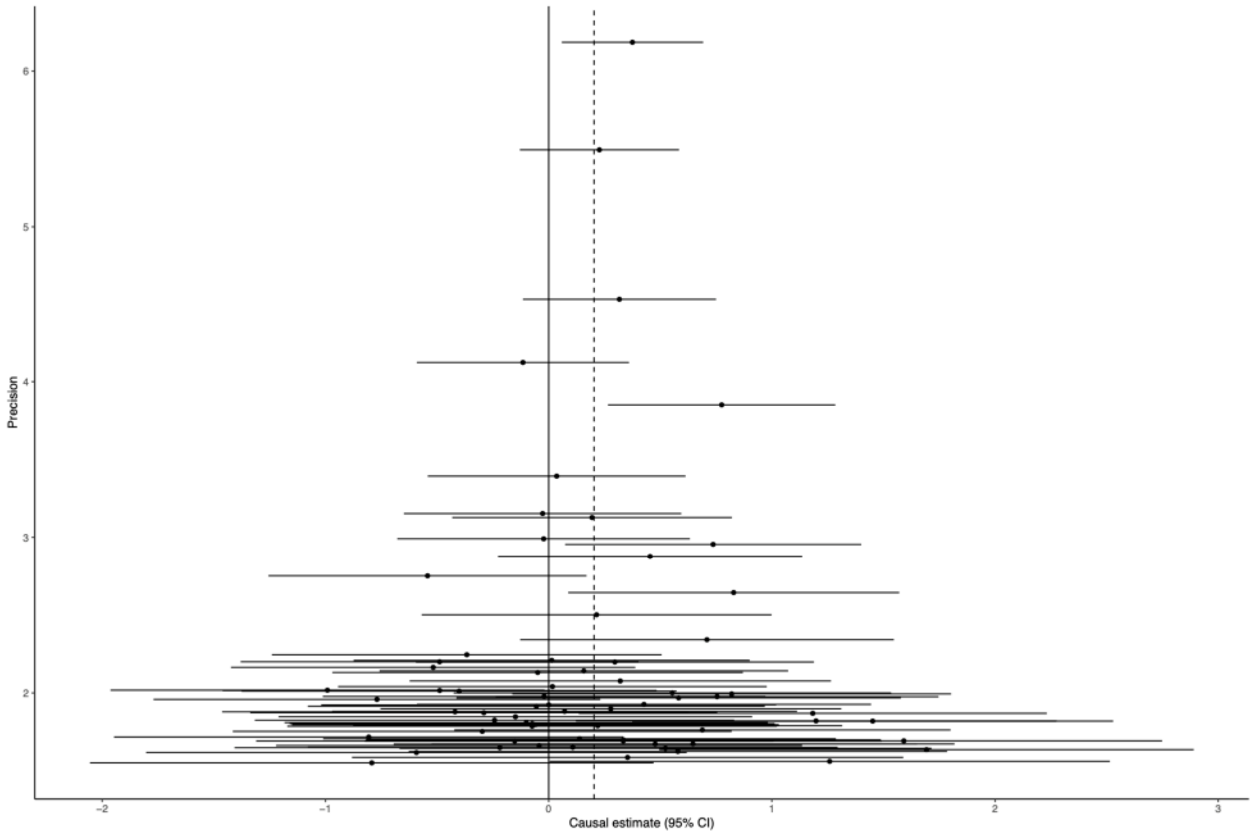


D) TSH

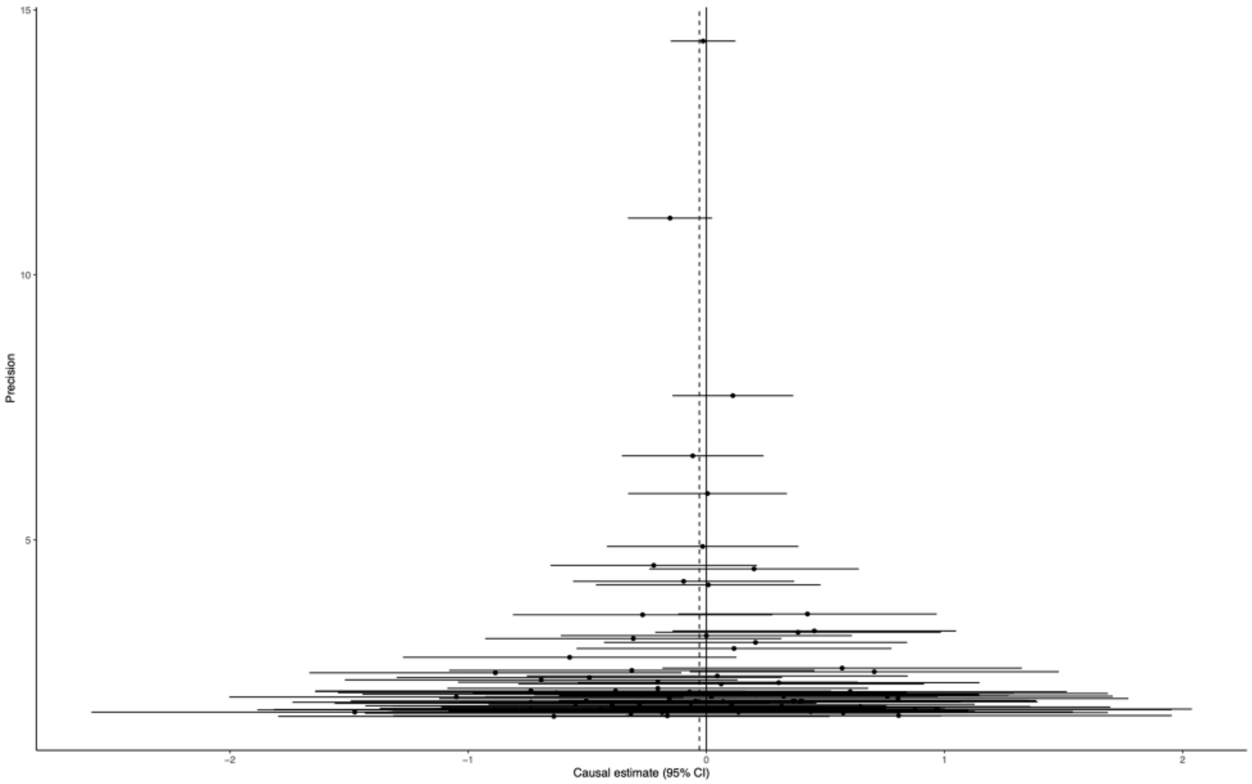

15
10
5
Precision
−2
−1
0
1
2
Causal estimate (95% CI)
